# Real World Evidence of the Neutralizing Monoclonal Antibody Sotrovimab for Preventing Hospitalization and Mortality in COVID-19 Outpatients

**DOI:** 10.1101/2022.04.03.22273360

**Authors:** Neil R. Aggarwal, Laurel E. Beaty, Tellen D. Bennett, Nichole E. Carlson, Christopher B. Davis, Bethany M. Kwan, David A. Mayer, Toan C. Ong, Seth Russell, Jeffrey Steele, Adane F. Wogu, Matthew K. Wynia, Richard D. Zane, Adit A. Ginde

**Affiliations:** Department of Medicine, University of Colorado School of Medicine, Aurora, 80045, USA; Department of Biostatistics and Informatics, Colorado School of Public Health, Aurora, 80045, USA; Section of Informatics and Data Science, Department of Pediatrics, University of Colorado School of Medicine, Aurora, 80045, USA; Department of Emergency Medicine, University of Colorado School of Medicine, Aurora, 80045, USA; Colorado Clinical and Translational Sciences Institute, University of Colorado Anschutz Medical Campus, Aurora, 80045, USA; Department of Family Medicine, University of Colorado School of Medicine, Aurora, 80045, USA; Research Informatics, Children’s Hospital Colorado, Aurora, 80045, USA; Center for Bioethics and Humanities, University of Colorado, Anschutz Medical Campus, Aurora, 80045, USA; Department of Health Systems Management and Policy, Colorado School of Public Health, Aurora, 80045, USA

**Keywords:** real-world evidence, COVID-19, sotrovimab, outpatients, mortality

## Abstract

**Background:** It is not known whether sotrovimab, a neutralizing monoclonal antibody (mAb) treatment authorized for early symptomatic COVID-19 patients, is effective against the SARS-CoV-2 Delta variant to prevent progression to severe disease and mortality.

**Methods:** Observational cohort study of non-hospitalized adult patients with SARS-CoV-2 infection from October 1^st^ 2021 - December 11^th^ 2021, using electronic health records from a statewide health system plus state-level vaccine and mortality data. We used propensity matching to select 3 patients not receiving mAbs for each patient who received outpatient sotrovimab treatment. The primary outcome was 28-day hospitalization; secondary outcomes included mortality and severity of hospitalization.

**Results:** Of 10,036 patients with SARS-CoV-2 infection, 522 receiving sotrovimab were matched to 1,563 not receiving mAbs. Compared to mAb-untreated patients, sotrovimab treatment was associated with a 63% decrease in the odds of all-cause hospitalization (raw rate 2.1% versus 5.7%; adjusted OR 0.37, 95% CI 0.19-0.66) and an 89% decrease in the odds of all-cause 28-day mortality (raw rate 0% versus 1.0%; adjusted OR 0.11, 95% CI 0.0-0.79), and may reduce respiratory disease severity among those hospitalized.

**Conclusion:** Real-world evidence demonstrated sotrovimab effectiveness in reducing hospitalization and all-cause 28-day mortality among COVID-19 outpatients during the Delta variant phase.

## BACKGROUND

High rates of transmission of severe acute respiratory syndrome coronavirus-2 (SARS-CoV-2), the virus that causes coronavirus disease of 2019 (COVID-19), persist, especially among unvaccinated individuals or those with waning vaccine or infection-related immunity.[1] Neutralizing monoclonal antibody (mAb) treatment provides immediate passive immunity against SARS-CoV-2. Several mAb products have received emergency use authorization (EUA) from the US Food and Drug Administration[2] based on Phase II/III randomized clinical trials conducted earlier in the pandemic that demonstrated efficacy towards reduced hospitalization and disease severity among high-risk outpatients.[3-5] Use of mAb products such as sotrovimab for individuals who have recently tested positive for SARS-CoV-2 in the outpatient setting is critical to mitigate virus-driven impact on the health care system and is also an evidence-based treatment strategy to improve COVID-19 outcomes among high-risk individuals.

Trials supporting mAb EUA approval were conducted prior to the emergence of the Delta variant surge in the summer 2021, and the trial assessing sotrovimab efficacy was limited in assessment of clinical outcomes or mortality.[4] Following EUA, however, it becomes more challenging to recruit patients into randomized controlled trials.[6] As new variants such as Delta and Omicron emerge, analysis of real-world data sufficiently robust to evaluate important clinical differences is critical to evaluate treatment effectiveness and inform policy and practice decisions. We previously used a real-world platform to report on mAb efficacy during the Delta variant pandemic phase.[7] That prior report focused on mAbs administered through September 2021, predominantly consisting of casivirimab plus imdevimab (∼80%), bamlanivimab (∼15%), or bamlanivimab plus etesevimab (∼3%). Sotrovimab had been administered to only 0.7% of the overall cohort of patients with the Delta variant. Yet, as of January 24^th^, 2022, sotrovimab is the only still-authorized mAb for outpatient COVID-19 treatment,[8] and thus additional data on its clinical effectiveness is warranted.

To provide useful data to help inform mAb allocation strategies and related policymaking, we leveraged our novel real-world evidence platform[7, 9, 10] to assess the clinical impact of sotrovimab therapy on high-risk outpatients with early symptomatic COVID-19 infections during a SARS-CoV-2 Delta predominant period in Colorado (10/1/2021 – 12/11/2021).[11] This paper reports on the effectiveness of sotrovimab against progression of COVID-19 to severe disease, hospitalization, severity of hospitalization, and death.

## METHODS

### Study Oversight and Data Sources

We conducted a propensity-matched observational cohort study, as part of a statewide implementation/effectiveness pragmatic trial, in a collaboration between University of Colorado researchers, University of Colorado Health (UCHealth) leaders, and the Colorado Department of Public Health and Environment (CDPHE). The study was approved by the Colorado Multiple Institutional Review Board with a waiver of informed consent. We obtained data from the electronic health record (EHR; Epic, Verona, WI) of UCHealth, the largest health system in Colorado with 13 hospitals around the state and 141,000 annual hospital admissions, using Health Data Compass, an enterprise-wide data warehouse. EHR data were merged with statewide data on vaccination status from the Colorado Comprehensive Immunization Information System and mortality from Colorado Vital Records.

### Patient Population Studied

We included patients diagnosed with SARS-CoV-2 infection between October 1, 2021 and December 11, 2021 allowing for at least 28 days of follow-up (n=10,036) (**Appendix Figure 1, Supplement**). Patients were identified using an EHR-based date of SARS-CoV-2 positive test (by polymerase chain reaction or antigen) or date of administration of mAb treatment (if no SARS-CoV-2 test result date available). The decision to seek mAb treatment was made by patients and clinicians.[11] We did not exclude patients solely for lack of EUA eligibility based on EHR data, because not all eligibility criteria were consistently available in the EHR. We excluded patients who tested positive for SARS-CoV-2 on the same day of or during hospitalization because they were not eligible for mAb treatment. We also excluded patients missing both a positive test date and a sotrovimab administration date (n=708), or if it had been more than 10 days between the positive test date and sotrovimab administration (n=26), resulting in a cohort of sotrovimab (N = 566) or mAb untreated (N = 9,470) patients.

Nearest neighbor propensity matching was conducted using logistic regression with treatment status as the outcome. Approximately three untreated patients (N=1,563) were matched to each sotrovimab-treated patient (N=522).[12, 13] Only 42 sotrovimab treated patients were lost due to incomplete covariate data. The propensity model included categorial age, sex, race/ethnicity, obesity status, immunocompromised status, number of comorbid conditions other than obesity and immunocompromised status, number of vaccinations at time of infection, and insurance status. We assessed effectiveness of matching using standardized mean differences (SMDs) with a threshold of 0.1 with results shown in Appendix Table 2.[14]

### Outcomes

The primary outcome was all-cause hospitalization within 28 days of a positive SARS-CoV-2 test, obtained from EHR data. Secondary outcomes included 28-day all-cause mortality, emergency department (ED) visit within 28 days, in-hospital disease severity based on maximum level of respiratory support, hospital and intensive care unit (ICU) lengths of stay (LOS) in survivors, rates of ICU admission, and in-hospital mortality. For both hospitalization and ED visits, the index visit was used. When mAb treated patients were missing a SARS-CoV-2 positive date (70.5%), we randomly imputed missing test dates from the distribution of observed time between SARS-CoV-2 to mAb administration.

### Variable Definitions

Hospitalization was defined as any inpatient or observation encounter documented in the EHR. ED visits were defined as any visit to the ED, with or without an associated inpatient or observation encounter. Presence of comorbid conditions and immunocompromised status were determined as reported previously[7] and described further in the Supplement. The number of comorbid conditions was calculated as the sum of the presence of hypertension, cardiovascular disease, diabetes, pulmonary disease, and renal disease.

COVID-19 disease severity was estimated using ordinal categories of respiratory support requirements at an encounter level, based on the highest level of support received among the following types (in increasing order): no supplemental oxygen, standard (nasal cannula/face mask) oxygen, high-flow nasal cannula or non-invasive ventilation, and invasive mechanical ventilation.[15] In-hospital mortality was the highest level of disease severity.

No virus sequencing results were available on an individual patient basis. However, this analysis focused on a period when the Delta variant was dominant (>99% by state-wide data)[11] and coupled with the timing of sotrovimab EUA and distribution in Colorado. Vaccination status was categorized by the number of vaccinations (0, 1, 2, or ≥ 3) administered prior to the date of the SARS-CoV-2 positive test.

The variables of interest include treatment status, categorical age in years, sex, race/ethnicity, insurance status, obesity status, immunocompromised status, number of additional comorbid conditions, and number of vaccinations. Due to small sample sizes, the variables age, race/ethnicity, insurance status, number of comorbid conditions, and vaccination status were each collapsed to the groups shown in the results tables.

### Statistical analysis

Firth’s logistic regression was used to assess the association between treatment and 28-day hospitalization, 28-day mortality, and 28-day ED visits. Firth’s logistic regression (R package logistf V 1.24) addresses estimation issues related to low event rates and complete separation.[16-18] All models were adjusted for age, sex, race/ethnicity, insurance status, obesity status, immunocompromised status, number of additional comorbid conditions, and number of vaccinations. The unadjusted number needed to treat (NNT) was calculated for hospitalization by treatment status.

Due to the small number of hospitalized participants, descriptive statistics including counts and raw rates were calculated for all secondary outcomes among hospitalized participants, including disease severity, hospital LOS, ICU visit, and ICU LOS.

Kaplan-Meier curves were estimated to visually assess cumulative incidence patterns by treatment status for 28-day hospitalization.

Two sensitivity analyses were performed. First, we repeated the above analysis using only EUA-eligible patients as verified by available EHR data. Second, we repeated the above analysis with a more conservative SARS-CoV-2 imputation approach where all missing positive test dates were imputed as ten days prior to the mAb administration date (the maximum time difference allowed by the EUA). All statistical analyses were performed using R Statistical Software (version 3.6.0; R Foundation for Statistical Computing, Vienna, Austria).[19]

## RESULTS

### Characteristics of sotrovimab-Treated and mAb-Untreated Cohorts

Of 10,036 patients with SARS-CoV-2 infection in the full cohort, 566 subjects received mAbs and 9,470 patients did not (**Appendix Table 1, Supplement**). In the full cohort, the sotrovimab-treated group generally reflects EUA criteria for use of mAbs, with many being older (33.0% were age ≥65 years vs. 11.4% in mAb-untreated group), more likely to be obese (24.0% vs. 16.8%), or having one or more comorbidities (49.1% vs. 36.5%). Propensity matching eliminated clinically meaningful differences in matching variables between groups (**Table 1, Appendix Table 2, Supplement**). 522 sotrovimab-treated patients were propensity matched to 1,563 untreated patients.

**Table 1.**
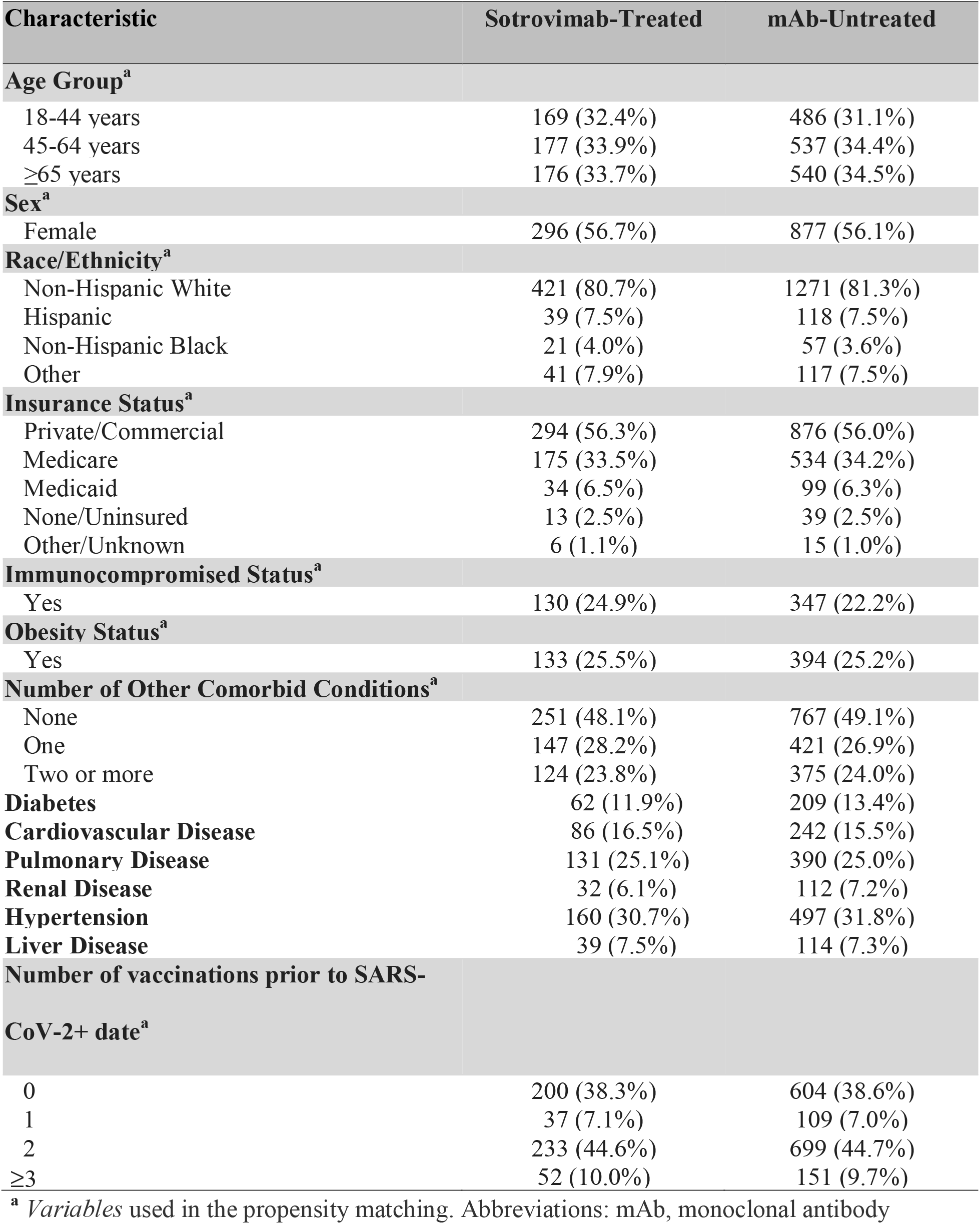
Baseline Characteristics by Mab Treatment Status for Primary Matched Cohort.

The characteristics of sotrovimab-treated and mAb-untreated patients in the matched cohort are presented (**Table 1**). The age distribution was similar, with 34% aged ε65. The cohort was 56% female, 81% Non-Hispanic white, and 56% had private/commercial insurance. Hypertension (32%) and pulmonary disease (25%) were the most common comorbid conditions. Notably, 54% had received at least two vaccinations at the time of infection, and 39% had not received any vaccine doses. The mean time from positive SARS-CoV-2 test to administration of sotrovimab treatment was 3.7 days (SD 1.8) in those who did not have an imputed positive test date.

### Hospitalization and Mortality

Sotrovimab treatment was associated with a lower rate of 28-day hospitalization compared to matched mAb-untreated controls (11 [2.1%] vs. 89 [5.7%]), representing a 63% decrease in the adjusted odds of hospitalization (adjusted OR 0.37, 95% CI 0.19-0.68; p < 0.001) (**Table 2**). The unadjusted NNT for hospitalization for the untreated group was 28. Based on a time-to-event analysis, the benefits associated with reduced hospitalization were largely accrued within 12 days of the positive SARS-CoV-2 test date (**Figure 1**), or an average of 9 days after sotrovimab treatment. Other factors associated with hospitalization are in Supplemental material (**Appendix Table 3, Supplement**). Covariates that were associated with increased odds of 28-day hospitalization included age ≥ 65 (p = 0.049), obesity (p < 0.001), and one (p = 0.013) or two or more (p < 0.001) comorbid conditions other than obesity or immunocompromised status (**Appendix Table 3, Supplement**). Each level of vaccination status (1, 2, or ≥ 3 doses) was significantly associated with decreased odds of hospitalization in comparison to having no vaccine.

**Table 2.** Primary and Secondary Outcomes by Monoclonal Antibody Treatment Status.

| Outcome | Sotrovimab-Treated | mAb-Untreated | Adjusted OR | 95% CI |
| --- | --- | --- | --- | --- |
| Overall Sample Size | N=522 | N=1563 |  |  |
| All-Cause 28-day Hospitalization (*primary outcome*) | 11 (2.1%) | 89 (5.7%) | 0.37 | (0.19, 0.68) |
| All-Cause 28-day Mortality | 0 (0.0%) | 15 (1.0%) | 0.11 | (0.00, 0.79) |
| Any ED visit to Day 28 | 44 (8.4%) | 119 (7.6%) | 1.12 | (0.77, 1.60) |
| Hospitalized Sample Size | N=11 | N=89 |  |  |
| Hospital LOS days, mean (SD) | 5.3 (5.9) | 9.4 (10.6) | -- | -- |
| IMV or Death | 0 (0.0%) | 19 (21.3%) | -- | -- |
| ICU Admission | 2 (18.2%) | 19 (21.3%) | -- | -- |
| ICU LOS days, mean (SD) | 5.5 (6.4) | 8.6 (10.1) | -- | -- |

All regression models adjusted for age, sex, race/ethnicity, obesity, immunocompromised status, number of comorbidities, insurance status, and vaccination status. Abbreviations: mAb, monoclonal antibody; OR, odds ratio; CI, confidence interval; ICU, intensive care unit; IMV, invasive mechanical ventilation; LOS, length of stay; SD, standard deviation

**Figure 1.**
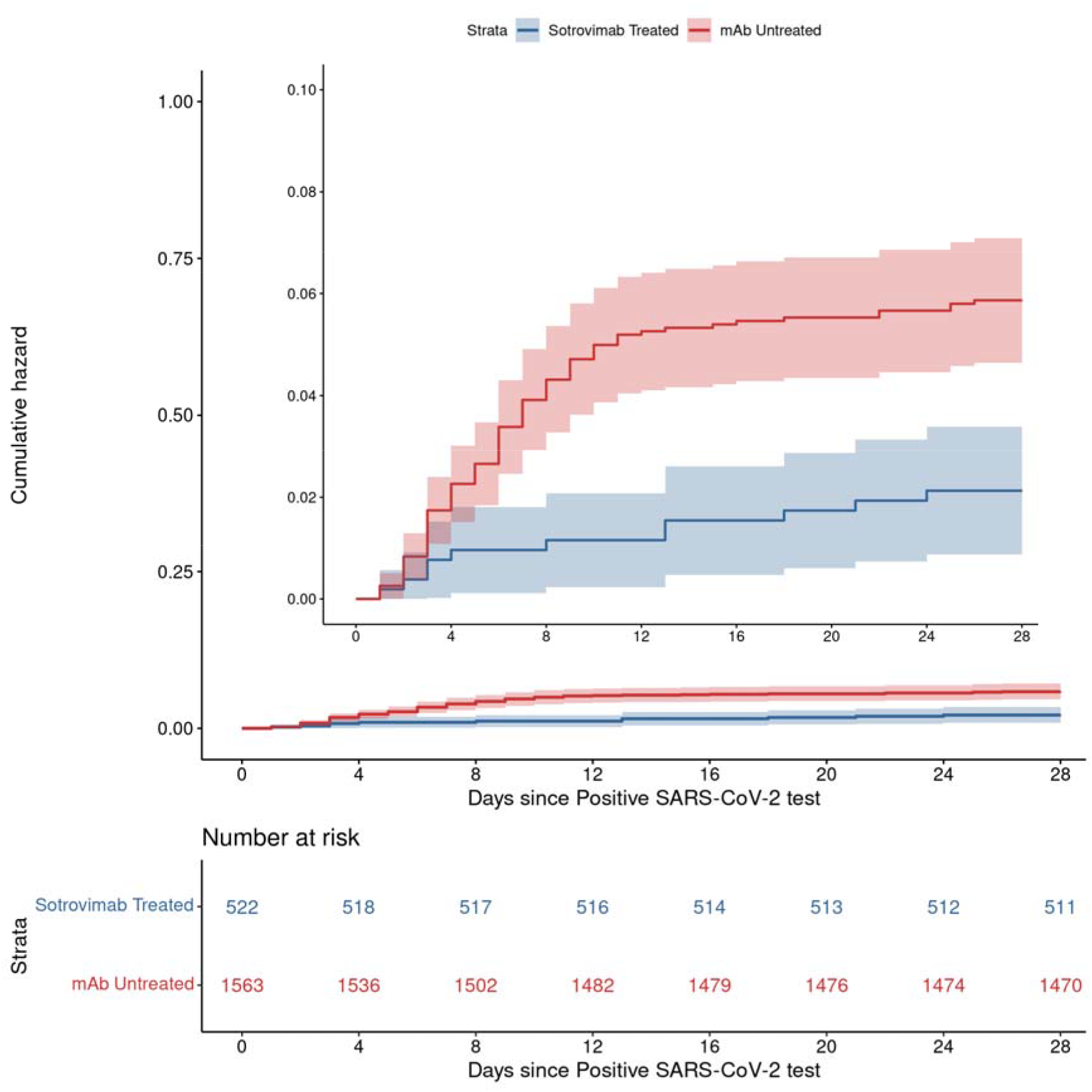
Cumulative Incidence Plots for All-Cause Hospitalization to Day 28 by Sotrovimab Treatment Status.

Importantly, all-cause 28-day mortality in the sotrovimab-treated group was 0 (0%) compared to 15 (1.0%) among the mAb-untreated group, equating to an 89% decrease in the mortality odds (adjusted OR 0.11, 95% CI 0.0-0.79 (**Table 2**). There was not a significant association between sotrovimab treatment and the odds of visiting the ED (adjusted OR 1.12, 95% CI 0.77-1.60).

### Severity of Hospitalization

Among hospitalized patients, 0 of 11 (0%) in the sotrovimab-treated group required invasive mechanical ventilation (IMV) or died in the hospital, compared to 19 of 89 (21.3%) mAb-untreated group (**Table 2**). We also observed that a higher proportion of sotrovimab-treated patients required no supplemental oxygen or only required standard (low-flow) oxygen in comparison to mAb-untreated patients (72.7% vs. 48.3%). The average hospital length of stay (LOS) for sotrovimab patients was 5.3 (+/- 5.9) days in comparison to 9.4 (+/- 10.6) days in the untreated group. Collectively, these data suggest a lower severity of disease among hospitalized sotrovimab-treated patients, although statistical inference was not performed (**Figure 2**). 2 of 9 (18.2%) sotrovimab-treated patients required ICU level of care, a similar percentage compared to mAb-untreated patients (21.3%).

**Figure 2.**
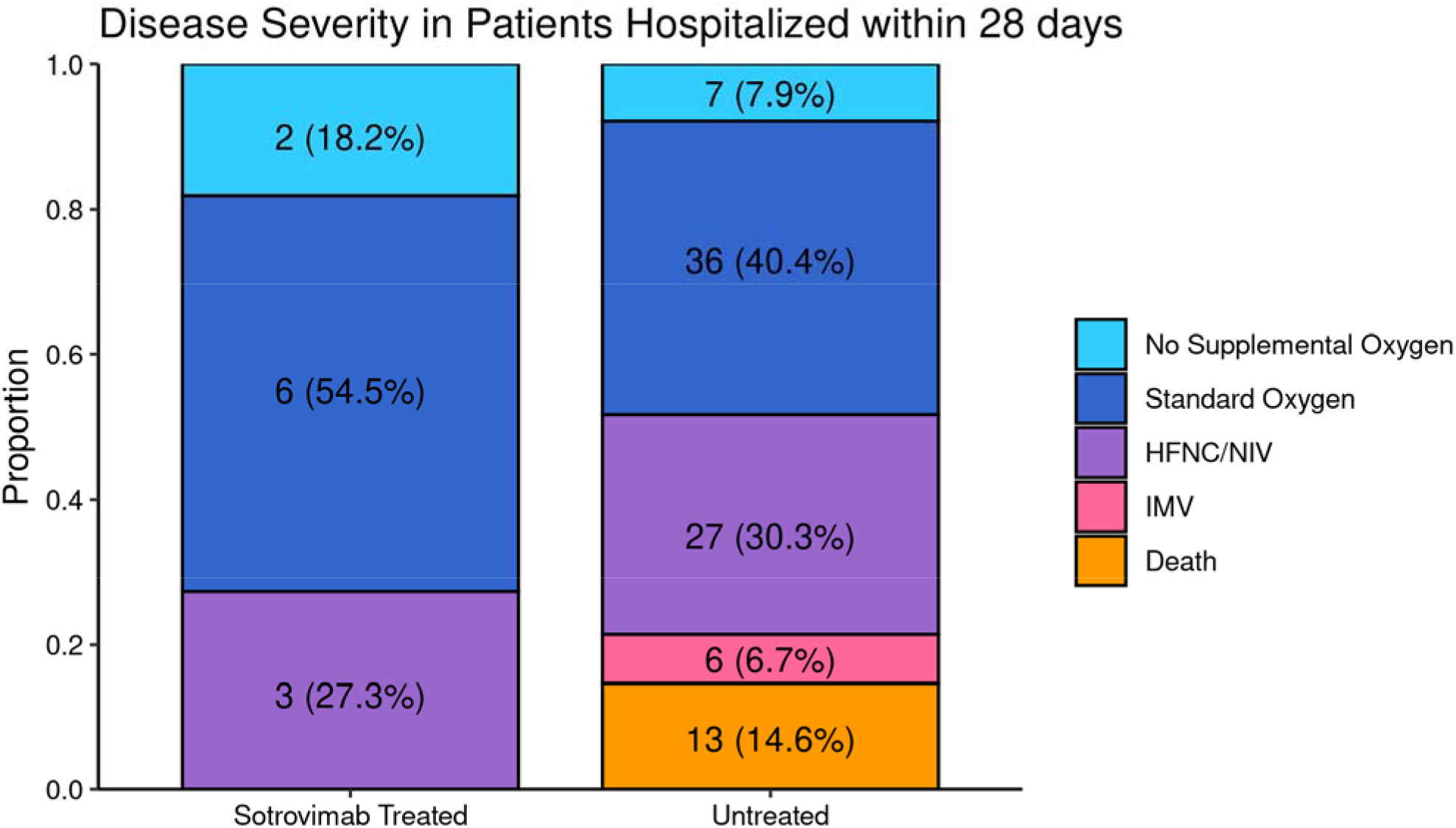
Maximum Respiratory Support by Monoclonal Antibody Treatment Status among Patients Hospitalized within 28 Days. Comparing severity of hospitalizations for n=11 sotrovimab-treated and n=89 mAb-untreated patients, the maximum level of respiratory support appeared lower for sotrovimab-treated patients, but inferential statistics were not able to be performed. Abbreviations: mAb, monoclonal antibody; HFNC, high-flow nasal cannula oxygen; NIV, non-invasive ventilation; IMV, invasive mechanical ventilation

### Sensitivity Analysis

Neither restricting the cohort to only patients meeting EUA eligibility criteria based on available EHR data or using a more conservative imputation method for missing date of positive SARS-CoV-2 test materially changed the key results and scientific conclusions (**Appendix Tables 4-7, Supplement**).

## DISCUSSION

During a SARS-CoV-2 Delta variant-predominant period in Colorado, sotrovimab reduced 28-day hospitalization by 63% and all-cause 28-day mortality by 89%. Our study adds to the main prior clinical trial demonstrating sotrovimab efficacy that had limited sample size (n=583), occurred prior to the emergence of the Delta variant, and was underpowered to evaluate the impact of sotrovimab on patient mortality.[4] This data on the effectiveness of sotrovimab to prevent severe COVID-19 disease induced by the Delta variant may be critical given the unpredictable nature of the COVID-19 pandemic. When combined with in vitro data that suggests sotrovimab effectively neutralizes the omicron variant,[20] these data support continued sotrovimab prioritization among outpatient treatment options.

Although we do not directly compare sotrovimab effectiveness to other mAbs used during a Delta variant-dominated pandemic phase, it is notable that the adjusted OR for 28-day hospitalization in our study (aOR 0.37) is similar to what we previously reported (aOR 0.48) when casivirimab plus imdevimab, bamlanivimab, or bamlanivimab plus etesevimab use as outpatient mAb therapy were far more common than sotrovimab.[7] Further supporting sotrovimab effectiveness against the Delta variant of SARS-CoV-2 is a comparable NNT of 28 to prevent one hospitalization, which we observed despite both an increase in the overall vaccination rate and a lower baseline hospitalization rate among mAb-untreated patients compared to the earlier cohort.[7]

Our results are of practical importance for policymakers and clinicians because there have been shortages of mAb supplies and infusion capacity, and as such, demonstrating their effectiveness to reduce hospitalization and mortality against each clinically relevant SARS-CoV-2 variant is crucial.[21, 22] Study findings support continued use of sotrovimab for patients with high baseline risk for hospitalization, including those who are older than 65, obese, are not fully vaccinated, or have comorbid conditions, all of whom had increased rates of hospitalization in our cohort.

### Limitations

This study has several limitations. The setting was a single health system and geographically limited to one US state with relatively low racial and ethnic minority representation, though it serves both urban and rural populations through academic and community hospitals. Even though we used statewide data for mortality and vaccination status, hospitalizations were collected only within one single health system. If mAb-untreated patients were less likely to be seen in this health system, hence more likely to be hospitalized elsewhere, this may bias our results toward the null. We also relied on EHR data, including manual chart reviews, which may have missing or inaccurate information about the presence of chronic conditions.[23] These factors might have limited our ability to detect the impact of sotrovimab treatment.

We only collected 28-day hospitalization and mortality data, and therefore we do not know whether sotrovimab effectiveness extends to a longer period after SARS-CoV-2 infection. However, our prior study would suggest that 28-day and 90-day data are similar with respect to hospitalization and mortality endpoints.[7] In this study, propensity scoring achieved excellent matching between mAb-treated and mAb-untreated patient groups across multiple variables, but unmeasured confounders may remain. Our EHR data does not contain information on SARS-CoV-2 variants at the patient level. However, during Colorado’s Delta phase more than 99% of sequenced SARS-CoV-2 was Delta variant.[11]

Finally, this study was conducted during a limited portion of the Delta variant-dominant period after sotrovimab drug distribution and infusion had been well-established in Colorado. In addition, hospitalization rates were lower over this same period, precluding our ability to perform inferential statistics on severity of illness among the hospitalized sub-cohort. Because the trend among the hospitalized subset strongly suggests sotrovimab benefit in reducing disease severity and hospital length of stay, examining sotrovimab effectiveness across US states during the same period would seemingly be an important next step to evaluate the generalizability of our findings. With the rapid emergence of the Omicron variant in mid-December 2021, our intent is to separately evaluate sotrovimab effectiveness against the omicron variant after enough treated and untreated cases have accrued. The Omicron variant has an abundance of mutations in the receptor-binding motif (RBM) of SARS-CoV-2, however in vitro reports suggest preserved neutralization of Omicron by sotrovimab,[24] potentially explained by its ability to target non-RBM epitopes shared across many sarbecoviruses, including SARS-CoV-1.

## Conclusion

This study demonstrated real-world evidence for effectiveness of sotrovimab treatment in reducing hospitalizations among COVID-19 outpatients during the Delta variant phase, as well as a remarkable 89% overall reduction in mortality at 28 days, compared to matched mAb-untreated patients. For hospitalized patients, prior outpatient sotrovimab treatment may reduce respiratory disease severity, hospital length of stay, and death, but a larger cohort is necessary to further examine this observation. When access to mAbs is limited, prioritizing patients at highest risk for hospitalization has the most potential to reduce health system strain during the COVID-19 pandemic.

## Data Availability

All data produced in the present study are available upon reasonable request to the authors

## FOOTNOTES

1. The authors do not have a commercial or other association that might pose a conflict of interest (e.g., pharmaceutical stock ownership, consultancy, advisory board membership, relevant patents, or research funding)
2. This study was funded by the National Center for Advancing Translational Sciences of the National Institutes of Health [grant numbers UL1TR002525, UL1TR002535-03S3 and UL1TR002535-04S2].
3. This work has not been presented in part or in entirety at any meetings

## Notes

### Competing Interest Statement

The authors have declared no competing interest.

### Author Declarations

The Colorado Multiple Institutional Review Board approved the study with a waiver of informed consent.

