## Supplementary Material for "Real World Evidence of the Neutralizing Monoclonal Antibody Sotrovimab for Preventing Hospitalization and Mortality in COVID-19 Outpatients"

This Appendix has been provided by the authors to give readers additional information about the work.

### Monoclonal Antibody (mAb) Colorado Research Team and Collaborators

University of Colorado Anschutz Medical Campus, Aurora, CO

***Project Leadership and Administration***: Adit A. Ginde, MD, MPH; Ronald J. Sokol, MD; Tim Lockie, MS, MBA; Heather R. Stocker, MA; Mujeeb Zaird, MBA

***Biostatistics, Epidemiology, and Research Design Core:*** Nichole E. Carlson, PhD; Laurel E. Beaty, MS; Adane F. Wogu, PhD; David A. Mayer, BS; Samantha Roberts, MPH; Megan Branda, MS

***Clinical Core***:; Matthew K. Wynia, MD, MPH; Neil Aggarwal, MD, MHSc; Julie G. Ressalam, MPH; Matthew DeCamp, MD, PHD**;** Hilary Lum, MD, PhD; Matthew A. Miller, PharmD, BCP, BCIDP; Kyle Molina, PharmD, BCIDP; Ruthvik Reddy Perikiti, MS; Lauren Shviraga Whitesitt, BS

***Informatics Core***: Tellen D. Bennett, MD, MS; Toan C. Ong, PhD; Seth Russell, MS; Jeffrey Steele, RN; Margaret Rebull, MA; Ian Brooks, PhD; Madelyne Hull, MPH; Brandy George; Aaron Mobley, PhD; Supported by the Health Data Compass Data Warehouse project team and Clinical Research Support Team (CReST)

***Dissemination and Implementation Core***: Bethany M. Kwan, PhD, MSPH; Vanessa Owen, MA; Chelsea Sobczak, MPH; Jenna Reno, PhD; Mika Hamer, MPH, PhD; Eric G. Campbell, PhD

University of Colorado Health (UCHealth) System

Richard D. Zane, MD; Christopher B. Davis, MD; Kathy Deanda, RN, MSN; Mathew Miller, PharmD; Kyle Molina, PharmD

Colorado Department of Public Health and Environment, Denver, CO

Eric France, MD, MSPH, MBA; Wendy M. Bamberg, MD; Alexis W. Burakoff, MD, MPH; Diana M. Tapay, MA; Shannon O’brien, MD, MPH; Amanda Hettinger, MA; Rachel Severson, MS; Elaine M. Sabyan

Tri-County Health Department

John M. Douglas, Jr, MD; Alix Rizzuto, MS, RN; Samantha Smith, MS, RDN

We also wish to thank patients and families for their participation in research to accelerate discovery and rapidly advance clinical care in the pandemic; numerous colleagues who provided support for this project; frontline health care workers for their tireless efforts and life-saving contributions; and researchers around the world working together in the quest to inform healthcare practice and improve patient outcomes of COVID-19.

### Supplementary Methods

##### Description of Demographic Variables

Age was determined at the time of positive SARS-CoV-2 test or mAb administration if SARS-CoV-2 test date was not available in the electronic health record (EHR). We categorized age into 18-44, 45-64, and ≥65 years, Sex was defined as legal sex in the EHR and was binarized into female and male (non-binary status was not explicitly defined), and this field was missing for two subjects. To preserve sample size, the variables race and ethnicity were combined and categorized into non-Hispanic white, non-Hispanic black, Hispanic, and other. The number of comorbid medical conditions was calculated using hypertension, cardiovascular disease, diabetes, pulmonary disease, and renal disease, and was categorized into none, one, or two or more. Immunocompromised status and obesity were categorized separately. All individual comorbid conditions were considered as binary variables with either evidence of the comorbid condition or no evidence of the comorbid condition in the EHR.

##### EHR Curation of Comorbidities

We defined comorbidities based on the updated Charlson and Elixhauser Comorbidity Indices[1, 2] as implemented in the ‘icd’ R package[3] and reported previously from the same health system.[4, 5] From the eligibility criteria above, we categorized a patient as having diabetes, renal disease, pulmonary disease, cardiovascular disease, or immunocompromised status if those conditions were present for that patient in either the Charlson or Elixhauser system. For obesity and hypertension, we used only the Elixhauser system. Because of the importance of immunocompromised status as a risk factor for hospitalization and mortality from COVID-19, we additionally defined patients as immunocompromised if any of the below medications were present in the EHR medication administration record during the 90-day lookback period. The list of medications was developed jointly by an expert team of UCHealth pharmacists and Infectious Disease physicians. Potentially discordant patients most often had received immune-suppressing medications prior to our IRB-approved 90-day lookback period or had received prednisone or methylprednisolone at doses under the expert-defined dose threshold.

*List of immune-suppressing medications*

- Alemtuzumab
- Belatacept in past 2 months
- Calcineurin inhibitors (tacrolimus and cyclosporine – excludes topical/ophthalmic administration routes)
- Eculizumab
- mTOR-inhibitors (everolimus, sirolimus – excludes topical routes)
- Mycophenolate, azathioprine, cyclophosphamide in the last 1 month
- Prednisone or methylprednisolone, oral or IV only (≥10 mg prednisone equivalent)
- Rituximab
- Thymoglobulin
- TNF-α inhibitor (e.g., infliximab, etanercept, golimumab, adalimumab, certolizumab)

##### Missing Data Techniques

Of the 566 patients who received sotrovimab treatment, 409 (72.3%) were missing an initial positive SARS-CoV-2 test date in the UCHealth EHR, suggesting many initial tests were performed outside the UCHealth system. After propensity matching, 368 (70.5%) had an imputed SARS-CoV-2 test date. For the primary analysis, a distribution of the time difference between positive SARS-CoV-2 test date and sotrovimab administration date was created for subjects who had both. Then, time differences were randomly sampled with replacement from this distribution and were used to impute positive test dates for the patients who only had a sotrovimab administration date. We evaluated a sensitivity analysis to this approach by imputing the maximum allowed time difference between SARS-CoV-2 positive date and sotrovimab administration date (10 days) for all patients missing the first date.

A complete case analysis was performed for propensity matching. All comorbid conditions were missing from the EHR for 624 (6.2%) patients. Race/ethnicity was missing for 406 (4.0%) of patients. A total of 892 (8.9%) of patients were removed prior to propensity matching for the complete case analysis. Although there are limitations to a complete case analysis, the ability to accurately impute missingness based on the available EHR variables was limited. Thus, the potential bias of a complete case analysis for this amount of missingness was believed to be smaller than the potential bias from a small imputation model.

##### Propensity Matching

The propensity matched dataset was created through a logistic regression propensity score matching process. Nearest neighbor matching was used, with a maximum ratio of 3:1 mAb-untreated and sotrovimab-treated groups. In the matching process, we lost few sotrovimab-treated and more mAb-untreated subjects and ended up with a ratio of approximately 3:1. A common support was used for both the cases and controls, and a caliper width of < 0.2*SD of the propensity distribution was applied.[6] The standardized mean differences (SMDs) of each level of all covariates included in the model were calculated to compare the means and prevalence in the propensity matched dataset. A SMD of <0.1 was considered to have a non-meaningful imbalance in the data.[7]

The baseline characteristics included in the propensity matching process were age in years, sex, race/ethnicity, immunocompromised status, obesity status, number of other comorbid conditions, and insurance status.

##### Model Fitting

Each of the models presented in Table 2 were fitted using the same group of adjustment variables. Univariate logistic regression models were fit with the primary outcome of all-cause hospitalization within the first 28 days after a SARS-CoV-2 positive test to assess the unadjusted association between each variable of interest and the outcome. The variables that were included were treatment status, categorical age in years, sex, race/ethnicity, insurance status, obesity status, immunocompromised status, number of comorbid conditions, and vaccination status. A significance level of 0.05 was used to determine statistical significance; 95% confidence intervals (CIs) were also used to evaluate clinical significance.

### Appendix Figures

##### **Appendix Figure 1: Flow of Patients into the Primary Study Cohort**

N = 16,198

Subject had a positive test prior to transition to Omicron variant (before 10/03/2021): N = 39,318

N = 16,906

56,224 subjects with a SARS CoV 2 positive test date on or before 12/11/21

N = 10,036

Subsetting to mAb untreated (N = 9,470) and Sotrovimab treated (566) only

N = 14,555

Subject had more than 10 days between positive test date and mAb administration date: N = 26

Propensity Matching

Missing both positive date and mAb administration date: N = 708

Propensity Matched Cohort

mAb-Treated: n = 2,675

Propensity Matched Cohort

mAb-Untreated: n = 6,677

N = 14,581

Hospitalization Criteria: Removed N = 1,617

1) Subject was admitted to hospital on the same day as their COVID + test (N = 124)
2) Subject was already admitted to the hospital at the time of their COVID + test (N = 1,493)

N = 1,563

N = 522

### Appendix Tables

##### Appendix Table 1. Baseline Characteristics by Monoclonal Antibody Treatment Status for Full SARS-CoV-2 Positive Cohort, Prior to Propensity Matching

| **Characteristic** | **mAb-Treated**  **n=566** | **mAb-Untreated n=9470** |
| --- | --- | --- |
| **Age Group *** |  |  |
| 18-44 years | 183 (32.3%) | 5578 (58.9%) |
| 45-64 years | 196 (34.6%) | 2815 (29.7%) |
| ≥65 years | 187 (33.0%) | 1077 (11.4%) |
| **Sex *** |  |  |
| Female | 319 (56.4%) | 5210 (55.0%) |
| **Race/Ethnicity** |  |  |
| Non-Hispanic White | 427 (75.4%) | 6360 (67.2%) |
| Hispanic | 39 (6.9%) | 1601 (16.9%) |
| Non-Hispanic Black | 21 (3.7%) | 492 (5.2%) |
| Other | 41 (7.2%) | 649 (6.9%) |
| Missing | 38 (6.7%) | 368 (3.9%) |
| **Insurance Status** |  |  |
| Private/Commercial | 323 (57.1%) | 6116 (64.6%) |
| Medicare | 183 (32.3%) | 1085 (11.5%) |
| Medicaid | 35 (6.2%) | 1420 (15.0%) |
| None/Uninsured | 19 (3.4%) | 520 (5.5%) |
| Other/Unknown | 6 (1.1%) | 329 (3.5%) |
| **Immunocompromised** |  |  |
| Yes | 133 (23.5%) | 1062 (11.2%) |
| Missing | 6 (1.1%) | 612 (6.5%) |
| **Obesity Status** |  |  |
| Yes | 136 (24.0%) | 1591 (16.8%) |
| Missing | 6 (1.1%) | 618 (6.5%) |
| **Number of Other Comorbid Conditions *** |  |  |
| 0 | 282 (49.8%) | 5390 (56.9%) |
| 1 | 153 (27.0%) | 2135 (22.5%) |
| ≥2 | 125 (22.1%) | 1327 (14.0%) |
| Missing | 6 (1.1%) | 618 (6.5%) |
| **Diabetes** |  |  |
| Yes | 63 (11.1%) | 750 (7.9%) |
| Missing | 6 (1.1%) | 618 (6.5%) |
| **Cardiovascular Disease** |  |  |
| Yes | 473 (83.6%) | 8149 (86.1%) |
| Missing | 6 (1.1%) | 618 (6.5%) |
| **Pulmonary Disease** |  |  |
| Yes | 136 (24.0%) | 1841 (19.4%) |
| Missing | 6 (1.1%) | 618 (6.5%) |
| **Renal Disease** |  |  |
| Yes | 32 (5.7%) | 360 (3.8%) |
| Missing | 6 (1.1%) | 618 (6.5%) |
| **Hypertension** |  |  |
| Yes | 1310 (47.5%) | 6321 (19.0%) |
| Missing | 6 (1.1%) | 618 (6.5%) |
| **Liver Disease** |  |  |
| Yes | 39 (6.9%) | 455 (4.8%) |
| Missing | 6 (1.1%) | 618 (6.5%) |
| **Number of Vaccinations** |  |  |
| 0 | 228 (40.3%) | 4780 (50.5%) |
| 1 | 40 (7.1%) | 701 (7.4%) |
| 2 | 243 (42.9%) | 3606 (38.1%) |
| 3+ | 55 (9.7%) | 383 (4.0%) |
| **Type of mAb Medication** |  |  |
| Not Applicable | 0 (0.0%) | 9470 (100.0%) |
| Sotrovimab | 566 (100.0%) | 0 (0.0%) |
| **Days to mAb Administration** |  |  |
| N missing | 0 | 9470 |
| Mean (SD) | 3.72 (1.77) | NA |
| Range | 0.000 - 10.000 | NA |

Abbreviations: mAb, monoclonal antibody; SD, standard deviation

##### Appendix Table 2. Standard Mean Differences in Matching Variables

| Matching Variable | Standard Mean Differences |
| --- | --- |
| Distance |  |
| Distance | 0.001 |
| Age in years |  |
| 18-44 | 0.029 |
| 45-64 | -0.008 |
| ≥65 | -0.020 |
| Sex |  |
| Female | 0.014 |
| Male | -0.014 |
| Race/Ethnicity |  |
| Non-Hispanic White | -0.018 |
| Hispanic | -0.002 |
| Non-Hispanic Black | 0.020 |
| Other | 0.014 |
| Obesity Status |  |
| Not obese | -0.005 |
| Obese | 0.005 |
| Immunocompromised Status |  |
| Not Immunocompromised | -0.059 |
| Immunocompromised | 0.059 |
| Number of Comorbid Conditions |  |
| 0 | -0.019 |
| 1 | 0.027 |
| ≥2 | -0.007 |
| Number of vaccinations prior to infection |  |
| 0 | -0.005 |
| 1 | 0.005 |
| 2 | 0.000 |
| ≥3 | 0.004 |
| Insurance Status |  |
| Private/Commercial | 0.008 |
| Medicare | -0.016 |
| Medicaid | 0.008 |
| None/Uninsured | 0.000 |
| Other/Unknown | 0.018 |

##### Appendix Table 3. Full Model Results for 28-Day Hospitalization Primary Outcome

| Characteristic | Adjusted OR | 95% CI | P-value |
| --- | --- | --- | --- |
| Treatment Status |  |  |  |
| mAb-Untreated | Reference |  |  |
| Sotrovimab-Treated | 0.37 | (0.19, 0.68) | <0.001 |
| Age Group |  |  |  |
| 18-44 | Reference |  |  |
| 45-64 | 1.51 | (0.75, 3.19) | 0.251 |
| ≥65 | 2.61 | (1.00, 7.03) | 0.049 |
| Sex |  |  |  |
| Female | Reference |  |  |
| Male | 1.15 | (0.74, 1.79) | 0.519 |
| Race/Ethnicity |  |  |  |
| Non-Hispanic White | Reference |  |  |
| Hispanic | 1.19 | (0.45, 2.71) | 0.704 |
| Non-Hispanic Black | 1.93 | (0.74, 4.51) | 0.17 |
| Other | 0.76 | (0.26, 1.81) | 0.56 |
| Insurance Status |  |  |  |
| Private/Commercial | Reference |  |  |
| Medicare | 1.54 | (0.71, 3.39) | 0.281 |
| Medicaid | 1.48 | (0.59, 3.37) | 0.387 |
| Other (None/Uninsured/Unknown) | 0.91 | (0.10, 3.95) | 0.917 |
| Obesity Status |  |  |  |
| No | Reference |  |  |
| Yes | 2.86 | (1.80, 4.57) | <0.001 |
| Immunocompromised Status |  |  |  |
| No | Reference |  |  |
| Yes | 1.05 | (0.65, 1.68) | 0.845 |
| Number of Comorbid Conditions |  |  |  |
| 0 | Reference |  |  |
| 1 | 2.24 | (1.18, 4.33) | 0.013 |
| ≥2 | 3.99 | (2.12, 7.76) | <0.001 |
| Vaccination Status (# of doses) |  |  |  |
| 0 | Reference |  |  |
| 1 | 0.37 | (0.13, 0.86) | 0.019 |
| 2 | 0.26 | (0.16, 0.42) | <0.001 |
| ≥3 | 0.22 | (0.08, 0.52) | <0.001 |

Abbreviations: mAb, monoclonal antibody; OR, odds ratio; CI, confidence interval

##### Appendix Table 4. Baseline Characteristics by Monoclonal Antibody Treatment Status for Sensitivity Analysis Cohort 1

| **Characteristic** | **Full Cohort** | | **Matched Cohort** | |
| --- | --- | --- | --- | --- |
|  | **Sotrovimab-Treated (n=459)** | **mAb-Untreated (n=6621)** | **Sotrovimab-Treated**  **(n = 440)** | **mAb-Untreated (n = 1318)** |
| **Age Group*** |  |  |  |  |
| 18-44 | 127 (27.7%) | 3522 (53.2%) | 125 (28.4%) | 355 (26.9%) |
| 45-64 | 145 (31.6%) | 2022 (30.5%) | 140 (31.8%) | 423 (32.1%) |
| ≥65 | 187 (40.7%) | 1077 (16.3%) | 175 (39.8%) | 540 (41.0%) |
| **Sex*** |  |  |  |  |
| Female | 266 (58.0%) | 3716 (56.1%) | 254 (57.7%) | 747 (56.7%) |
| **Race/Ethnicity*** |  |  |  |  |
| Non-Hispanic White | 343 (74.7%) | 3800 (57.4% | 339 (77.0%) | 1027 (77.9%) |
| Hispanic | 39 (8.5%) | 1601 (24.2%) | 39 (8.9%) | 124 (9.4%) |
| Non-Hispanic Black | 21 (4.6%) | 492 (7.4%) | 21 (4.8%) | 50 (3.8%) |
| Other | 41 (8.9%) | 649 (9.8%) | 41 (9.3%) | 117 (8.9%) |
| Missing | 15 (3.3%) | 79 (1.2%) | -- | -- |
| **Insurance Status*** |  |  |  |  |
| Private/Commercial | 235 (51.2%) | 3861 (58.3%) | 226 (51.4%) | 684 (51.9%) |
| Medicare | 183 (39.9%) | 1071 (16.2%) | 174 (39.5%) | 519 (39.4%) |
| Medicaid | 28 (6.1%) | 1148 (17.3%) | 28 (6.4%) | 88 (6.7%) |
| None/Uninsured | 10 (2.2%) | 312 (4.7%) | 9 (2.0%) | 26 (2.0%) |
| Other/Unknown | 3 (0.7%) | 229 (3.5%) | 3 (0.7%) | 1 (0.1%) |
| **Obesity Status*** |  |  |  |  |
| Yes | 136 (29.6%) | 1591 (24.0%) | 134 (30.5%) | 389 (29.5%) |
| Missing | 2 (0.4%) | 242 (3.7%) | -- | -- |
| **Immunocompromised*** |  |  |  |  |
| Yes | 133 (29.0%) | 1062 (16.0%) | 129 (29.3%) | 374 (28.4%) |
| Missing | 2 (0.4%) | 236 (3.6%) | -- | -- |
| **Number of Other Comorbid Conditions*** |  |  |  |  |
| None | 179 (39.0%) | 2917 (44.1%) | 168 (38.2%) | 527 (40.0%) |
| One | 153 (33.3%) | 2135 (32.2%) | 147 (33.4%) | 400 (30.3%) |
| Two or more | 125 (27.2%) | 1327 (20.0%) | 125 (28.4%) | 391 (29.7%) |
| Missing | 2 (0.4%) | 242 (3.7%) | -- | -- |
| **Diabetes Status** |  |  |  |  |
| Yes | 63 (13.7%) | 750 (11.3%) | 63 (14.3%) | 199 (15.1%) |
| Missing | 2 (0.4%) | 242 (3.7%) | -- | -- |
| **Cardiovascular Disease Status** |  |  |  |  |
| Yes | 87 (19.0%) | 703 (10.6%) | 87 (19.8%) | 255 (19.3%) |
| Missing | 2 (0.4%) | 242 (3.7%) | -- | -- |
| **Pulmonary Disease** |  |  |  |  |
| Yes | 136 (29.6%) | 1841 (27.8%) | 132 (30.0%) | 389 (29.5%) |
| Missing | 2 (0.4%) | 242 (3.7%) | -- | -- |
| **Renal Disease** |  |  |  |  |
| Yes | 32 (7.0%) | 360 (5.4%) | 32 (7.3%) | 114 (8.6%) |
| Missing | 2 (0.4%) | 242 (3.7%) | -- | -- |
| **Hypertension** |  |  |  |  |
| Yes | 162 (35.3%) | 1923 (29.0%) | 160 (36.4%) | 495 (37.6%) |
| Missing | 2 (0.4%) | 242 (3.7%) | -- | -- |
| **Liver Disease Status** |  |  |  |  |
| Yes | 38 (8.3%) | 427 (6.5%) | 38 (8.6%) | 108 (8.2%) |
| Missing | 2 (0.4%) | 242 (3.7%) | -- | -- |
| **Number of vaccinations prior to SARS-CoV-2+ date*** |  |  |  |  |
| 0 | 173 (37.7%) | 3387 (51.2%) | 163 (37.0%) | 494 (37.5%) |
| 1 | 34 (7.4%) | 477 (7.2%) | 32 (7.3%) | 83 (6.3%) |
| 2 | 201 (43.8%) | 2473 (37.4%) | 197 (44.8%) | 615 (46.7%) |
| 3+ | 51 (11.1%) | 284 (4.3%) | 48 (10.9%) | 126 (9.6%) |
| **Type of mAb Administration** |  |  |  |  |
| Not Applicable | 0 (0.0%) | 6621 (100.0%) | 0 (0.0%) | 1318 (100.0%) |
| Sotrovimab | 459 (100.0%) | 0 (0.0%) | 440 (100.0%) | 0 (0.0%) |

Abbreviations: mAb, monoclonal antibody; SD, standard deviation

##### Appendix Table 5. Model Comparison for Sensitivity Analysis Cohort 1 for 28-day hospitalization primary outcome

|  | Full Matched Model | | | Eligible Matched Model | | | |
| --- | --- | --- | --- | --- | --- | --- | --- |
|  | OR | 95% CI | P-value | OR | 95% CI | | P-value |
| Treatment Status |  |  |  |  | |  |  |
| mAb-Untreated | Reference |  |  |  | |  |  |
| Sotrovimab-Treated | 0.37 | (0.19, 0.68) | <0.001 | 0.39 | | (0.19, 0.71) | 0.001 |
| Age Group |  |  |  |  | |  |  |
| 18-44 | Reference |  |  |  | |  |  |
| 45-64 | 1.51 | (0.75, 3.19) | 0.251 | 1.37 | | (0.65, 3.04) | 0.407 |
| ≥65 | 2.61 | (1.00, 7.03) | 0.049 | 2.8 | | (0.99, 8.14) | 0.053 |
| Sex |  |  |  |  | |  |  |
| Female | Reference |  |  |  | |  |  |
| Male | 1.15 | (0.74, 1.79) | 0.519 | 0.98 | | (0.62, 1.54) | 0.921 |
| Race/Ethnicity |  |  |  |  | |  |  |
| Non-Hispanic White | Reference |  |  |  | |  |  |
| Hispanic | 1.19 | (0.45, 2.71) | 0.704 | 0.45 | | (0.12, 1.25) | 0.135 |
| Non-Hispanic Black | 1.93 | (0.74, 4.51) | 0.17 | 1.16 | | (0.35, 3.07) | 0.794 |
| Other | 0.76 | (0.26, 1.81) | 0.56 | 0.8 | | (0.30, 1.83) | 0.616 |
| Insurance Status |  |  |  |  | |  |  |
| Private/Commercial | Reference |  |  |  | |  |  |
| Medicare | 1.54 | (0.71, 3.39) | 0.281 | 1.59 | | (0.68, 3.84) | 0.286 |
| Medicaid | 1.48 | (0.59, 3.37) | 0.387 | 2.09 | | (0.85, 4.85) | 0.106 |
| Other (None/Uninsured/Unknown) | 0.91 | (0.10, 3.95) | 0.917 | 1.92 | | (0.20, 8.61) | 0.499 |
| Obesity Status |  |  |  |  | |  |  |
| No | Reference |  |  |  | |  |  |
| Yes | 2.86 | (1.80, 4.57) | <0.001 | 2.82 | | (1.78, 4.49) | <0.001 |
| Immunocompromised Status |  |  |  |  | |  |  |
| No | Reference |  |  |  | |  |  |
| Yes | 1.05 | (0.65, 1.68) | 0.845 | 1.2 | | (0.74, 1.95) | 0.452 |
| Number of Comorbid Conditions |  |  |  |  | |  |  |
| 0 | Reference |  |  |  | |  |  |
| 1 | 2.24 | (1.18, 4.33) | 0.013 | 1.86 | | (0.97, 3.67) | 0.062 |
| ≥2 | 3.99 | (2.12, 7.76) | <0.001 | 2.93 | | (1.54, 5.76) | <0.001 |
| Vaccination Status (# of doses) |  |  |  |  | |  |  |
| 0 | Reference |  |  |  | |  |  |
| 1 | 0.37 | (0.13, 0.86) | 0.019 | 0.34 | | (0.11, 0.84) | 0.017 |
| 2 | 0.26 | (0.16, 0.42) | <0.001 | 0.24 | | (0.15, 0.40) | <0.001 |
| ≥3 | 0.22 | (0.08, 0.52) | <0.001 | 0.16 | | (0.04, 0.41) | <0.001 |

Abbreviations: mAb, monoclonal antibody; OR, odds ratio; CI, confidence interval; LOS, length of stay; ICU, intensive care unit; SD, standard deviation

**Appendix Table 6. Baseline Characteristics by Monoclonal Antibody Treatment Status for Sensitivity Analysis Cohort 2**

|  | **Full Cohort** | | **Matched Cohort** | |
| --- | --- | --- | --- | --- |
|  | **Sotrovimab-Treated  (n=610)** | **mAb-Untreated  (n=9470)** | **Sotrovimab-Treated  (n=560)** | **mAb-Untreated  (n=1680)** |
| **Age Group*** |  |  |  |  |
| 18-44 years | 199 (32.6%) | 5578 (58.9%) | 183 (32.7%) | 522 (31.1%) |
| 45-64 years | 217 (35.6%) | 2815 (29.7%) | 198 (35.4%) | 581 (34.6%) |
| ≥65 years | 194 (31.8%) | 1077 (11.4%) | 179 (32.0%) | 577 (34.3%) |
| **Sex*** |  |  |  |  |
| Female | 350 (57.4%) | 5210 (55.0%) | 323 (57.7%) | 939 (55.9%) |
| **Race/Ethnicity*** |  |  |  |  |
| Non-Hispanic White | 462 (75.7%) | 6360 (67.2%) | 452 (80.7%) | 1379 (82.1%) |
| Hispanic | 41 (6.7%) | 1601 (16.9%) | 41 (7.3%) | 127 (7.6%) |
| Non-Hispanic Black | 22 (3.6%) | 492 (5.2%) | 22 (3.9%) | 54 (3.2%) |
| Other | 45 (7.4%) | 649 (6.9%) | 45 (8.0%) | 120 (7.1%) |
| Missing | 40 (6.6%) | 368 (3.9%) | -- | -- |
| **Insurance Status*** |  |  |  |  |
| Private/Commercial | 350 (57.4%) | 6116 (64.6%) | 319 (57.0%) | 957 (57.0%) |
| Medicare | 186 (30.5%) | 1085 (11.5%) | 175 (31.2%) | 540 (32.1%) |
| Medicaid | 44 (7.2%) | 1420 (15.0%) | 43 (7.7%) | 128 (7.6%) |
| None/Uninsured | 22 (3.6%) | 520 (5.5%) | 15 (2.7%) | 42 (2.5%) |
| Other/Unknown | 8 (1.3%) | 329 (3.5%) | 8 (1.4%) | 13 (0.8%) |
| **Obesity Status*** |  |  |  |  |
| Yes | 147 (24.1%) | 1591 (16.8%) | 144 (25.7%) | 413 (24.6%) |
| Missing | 7 (1.1%) | 618 (6.5%) | -- | -- |
| **Immunocompromised Status*** |  |  |  |  |
| Yes | 138 (22.6%) | 1062 (11.2%) | 132 (23.6%) | 345 (20.5%) |
| Missing | 7 (1.1%) | 612 (6.5%) | -- | -- |
| **Number of Other Comorbid Conditions*** |  |  |  |  |
| None | 312 (51.1%) | 5390 (56.9%) | 276 (49.3%) | 842 (50.1%) |
| One | 161 (26.4%) | 2135 (22.5%) | 155 (27.7%) | 458 (27.3%) |
| Two or more | 130 (21.3%) | 1327 (14.0%) | 129 (23.0%) | 380 (22.6%) |
| Missing | 7 (1.1%) | 618 (6.5%) | -- | -- |
| **Diabetes Status** |  |  |  |  |
| Yes | 66 (10.8%) | 750 (7.9%) | 65 (11.6%) | 202 (12.0%) |
| Missing | 7 (1.1%) | 618 (6.5%) | -- | -- |
| **Cardiovascular Disease Status** |  |  |  |  |
| Yes | 90 (14.8%) | 703 (7.4%) | 89 (15.9%) | 247 (14.7%) |
| Missing | 7 (1.1%) | 618 (6.5%) | -- | -- |
| **Pulmonary Disease Status** |  |  |  |  |
| Yes | 143 (23.4%) | 1841 (19.4%) | 138 (24.6%) | 405 (24.1%) |
| Missing | 7 (1.1%) | 618 (6.5%) | -- | -- |
| **Renal Disease Status** |  |  |  |  |
| Yes | 33 (5.4%) | 360 (3.8%) | 33 (5.9%) | 115 (6.8%) |
| Missing | 7 (1.1%) | 618 (6.5%) | -- | -- |
| **Hypertension Status** |  |  |  |  |
| Yes | 169 (27.7%) | 1923 (20.3%) | 167 (29.8%) | 513 (30.5%) |
| Missing | 7 (1.1%) | 618 (6.5%) | -- | -- |
| **Liver Disease Status** |  |  |  |  |
| Yes | 41 (6.7%) | 455 (4.8%) | 41 (7.3%) | 114 (6.8%) |
| Missing | 7 (1.1%) | 618 (6.5%) | -- | -- |
| **Number of vaccinations prior to SARS-CoV-2+ date** |  |  |  |  |
| 0 | 252 (41.3%) | 4780 (50.5%) | 221 (39.5%) | 683 (40.7%) |
| 1 | 42 (6.9%) | 701 (7.4%) | 40 (7.1%) | 104 (6.2%) |
| 2 | 262 (43.0%) | 3606 (38.1%) | 251 (44.8%) | 773 (46.0%) |
| 3+ | 54 (8.9%) | 383 (4.0%) | 48 (8.6%) | 120 (7.1%) |
| **Type of mAb Administration** |  |  |  |  |
| Not Applicable | 0 (0.0%) | 9470 (100.0%) | 0 (0.0%) | 1680 (100.0%) |
| Sotrovimab | 610 (100.0%) | 0 (0.0%) | 560 (100.0%) | 0 (0.0%) |

Abbreviations: mAb, monoclonal antibody, SD, standard deviation

##### Appendix Table 7. Model Comparison for Sensitivity Analysis Cohort 2 for 28-day hospitalization primary outcome

|  | Full Matched Model | | | Eligible Matched Model | | | |
| --- | --- | --- | --- | --- | --- | --- | --- |
|  | OR | 95% CI | P-value | OR | 95% CI | | P-value |
| Treatment Status |  |  |  |  | |  |  |
| mAb-Untreated | Reference |  |  |  | |  |  |
| Sotrovimab Treated | 0.37 | (0.19, 0.68) | <0.001 | 0.32 | | (0.16, 0.60) | <0.001 |
| Age in years |  |  |  |  | |  |  |
| 18-54 | Reference |  |  |  | |  |  |
| 45-64 | 1.51 | (0.75, 3.19) | 0.251 | 1.29 | | (0.64, 2.73) | 0.49 |
| ≥65 | 2.61 | (1.00, 7.03) | 0.049 | 3.48 | | (1.43, 8.63) | 0.006 |
| Sex |  |  |  |  | |  |  |
| Female | Reference |  |  |  | |  |  |
| Male | 1.15 | (0.74, 1.79) | 0.519 | 1.1 | | (0.71, 1.71) | 0.666 |
| Race/Ethnicity |  |  |  |  | |  |  |
| Non-Hispanic White | Reference |  |  |  | |  |  |
| Hispanic | 1.19 | (0.45, 2.71) | 0.704 | 1.03 | | (0.35, 2.53) | 0.946 |
| Non-Hispanic Black | 1.93 | (0.74, 4.51) | 0.17 | 1.61 | | (0.52, 4.19) | 0.38 |
| Other | 0.76 | (0.26, 1.81) | 0.56 | 1.34 | | (0.57, 2.84) | 0.482 |
| Insurance Status |  |  |  |  | |  |  |
| Private/Commercial | Reference |  |  |  | |  |  |
| Medicare | 1.54 | (0.71, 3.39) | 0.281 | 1.08 | | (0.54, 2.20) | 0.825 |
| Medicaid | 1.48 | (0.59, 3.37) | 0.387 | 0.9 | | (0.35, 2.08) | 0.818 |
| Other (None/Uninsured/Unknown) | 0.91 | (0.10, 3.95) | 0.917 | 0.6 | | (0.06, 2.51) | 0.532 |
| Obesity Status |  |  |  |  | |  |  |
| No | Reference |  |  |  | |  |  |
| Yes | 2.86 | (1.80, 4.57) | <0.001 | 3.28 | | (2.07, 5.21) | <0.001 |
| Immunocompromised Status |  |  |  |  | |  |  |
| No | Reference |  |  |  | |  |  |
| Yes | 1.05 | (0.65, 1.68) | 0.845 | 0.92 | | (0.56, 1.47) | 0.719 |
| Number of Comorbid Conditions |  |  |  |  | |  |  |
| 0 | Reference |  |  |  | |  |  |
| 1 | 2.24 | (1.18, 4.33) | 0.013 | 1.99 | | (1.05, 3.83) | 0.036 |
| ≥2 | 3.99 | (2.12, 7.76) | <0.001 | 5.36 | | (2.90, 10.23) | <0.001 |
| Number of vaccinations prior to SARS-CoV-2+ date |  |  |  |  | |  |  |
| 0 | Reference |  |  |  | |  |  |
| 1 | 0.37 | (0.13, 0.86) | 0.019 | 0.33 | | (0.12, 0.79) | 0.011 |
| 2 | 0.26 | (0.16, 0.42) | <0.001 | 0.18 | | (0.11, 0.29) | <0.001 |
| ≥3 | 0.22 | (0.08, 0.52) | <0.001 | 0.17 | | (0.04, 0.45) | <0.001 |

Abbreviations: mAb, monoclonal antibody; OR, odds ratio; CI, confidence interval; SD, standard deviation

##### Appendix Table 8. Monoclonal Antibody Emergency Use Authorization Eligibility Criteria

| Prior to June 1, 2021 | After June 1, 2021 |
| --- | --- |
| Body mass index of 35 kg/m^2^ or more | Body mass index of 25 kg/m^2^ or more |
| Chronic kidney disease | Chronic kidney disease |
| Diabetes | Diabetes |
| Immunosuppressive disease or currently receiving immunosuppressive treatment | Immunosuppressive disease or are currently receiving immunosuppressive treatment |
| 65 years of age or older | 65 years of age or older |
| 55 years of age or older AND have either cardiovascular disease OR hypertension OR chronic obstructive pulmonary disease/other chronic respiratory disease | Chronic respiratory diseases, cardiovascular disease, or hypertension |
|  | Pregnancy |
|  | Sickle cell disease |
|  | Neurodevelopmental disorders |
|  | Medical related technology dependence |
|  | Other medical conditions or factors (e.g., race or ethnicity) that places individual patients at risk for progression to severe COVID-19 |
